# Default Handling of the Non-Assessable Verbal Glasgow Coma Scale Misclassifies Illness Severity in Mechanically Ventilated Patients: A Retrospective Analysis

**DOI:** 10.64898/2026.06.20.26356135

**Authors:** Alon Gorenshtein, Yosef Adiniaev, Mahmud Omar, Yiftach Barash, Eyal Klang, Oved Daniel

## Abstract

**Background:** The Glasgow Coma Scale (GCS) is the intensive care unit’s universal neurologic severity score and is embedded in the Acute Physiology and Chronic Health Evaluation (APACHE) and Sequential Organ Failure Assessment (SOFA), risk-adjusted mortality models, and ICU benchmarking. In the mechanically ventilated patient the verbal component cannot be obtained, and the conventions used to fill that gap (defaulting the total to a normal score of 15, or dropping the patient from the analysis) may relabel the sickest, most-sedated patients as neurologically normal and distort illness-severity estimates.

**Objective:** To quantify non-assessable GCS verbal examinations after acute brain injury and how the handling convention changes severity scoring and mortality-model behavior, in two independent critical care databases.

**Materials and Methods:** Retrospective cohort of adults with acute brain injury, first intensive care unit stay, MIMIC-IV. A verbal examination was non-assessable when documented “No Response-ETT.” We quantified burden and determinants, compared the MIMIC-IV derived GCS convention, which defaults intubated patients to 15, with a component-aware GCS, and audited mortality-model handling strategies.

**Results:** Among 14,230 patients, 45.2% had a non-assessable verbal examination; 47.5% of ventilated patients had none in the first 24 hours. Non-assessability was driven by mechanical ventilation (odds ratio, 27.4; 95% CI, 22.8 to 33.0) and associated with death (odds ratio, 1.56; 95% CI, 1.32 to 1.85). The MIMIC-IV derived GCS was 15 in 42.9% of patients, placed 11.6% in the lowest severity category despite eye and motor indicating GCS 9 or less, and discriminated mortality worse than a component-aware GCS (AUROC, 0.746 vs 0.783). Complete-case handling excluded 28.5% of patients with 50.2% of deaths. Both the default-to-normal convention and the complete-case selection trap replicated in the multi-hospital eICU-CRD database (among intubated stays, 25.2% carried a total GCS of 15; non-assessable stays had higher mortality than assessable stays, 16.6% vs 9.2%).

**Discussion:** A derived-score convention can make the sickest intubated patients appear neurologically normal, while complete-case handling removes the highest-risk patients from analysis.

**Conclusion:** In mechanically ventilated patients the default handling of the non-assessable verbal GCS misclassifies illness severity, and this distortion replicated across two independent databases including 171 hospitals in eICU-CRD/APACHE, so it is a general property of ICU scoring rather than a single-database quirk. ICU severity scores, risk-adjusted mortality models, and GCS-based benchmarking or quality metrics should flag and report how the non-assessable verbal examination was handled.

## Introduction

The Glasgow Coma Scale (GCS) is the intensive care unit’s universal neurologic severity score and the principal real-time signal of neurological change at the bedside^1,2^. It is also embedded in the severity scores and risk-adjustment systems that govern critical care: it contributes to the Acute Physiology and Chronic Health Evaluation (APACHE) and to the central nervous system component of the Sequential Organ Failure Assessment (SOFA), it feeds risk-adjusted mortality models and ICU benchmarking, and it informs bedside decisions^12,13^. The scale combines an eye-opening, a verbal, and a motor response, recorded by nurses many times each day. Two features of critical illness, however, remove the examination from view. The verbal component cannot be obtained from an intubated patient, and continuous sedation, which is administered to most mechanically ventilated brain-injured patients, suppresses all three components; neuromuscular blockade abolishes the motor response entirely^3,4^. These are precisely the conditions under which the examination is most needed and least available.

When the verbal component cannot be obtained, a total score must still be produced, and the conventions used to fill the gap differ across critical care practice and research: discarding the affected examinations, imputing a fixed value, reporting an eye-plus-motor sum, or defaulting the total to a normal score of 15. Methods exist to estimate a total score, most prominently the eye-and-motor-based imputation of Brennan and colleagues, which has been validated for intubated patients with traumatic brain injury in a single-center database^5,6^. These methods address how to fill the gap within one population. They do not describe how large the gap is across the range of acute brain injuries, when during the critical illness it opens and closes, or what happens to an illness-severity score or a risk-adjusted mortality model when the gap is handled by these differing conventions^7,16,17^.

Because the examination disappears for reasons tied to severity and treatment rather than at random, its absence is informative, and the convention chosen to handle it may silently relabel the sickest, most-sedated patients as neurologically normal.

Whether non-assessable verbal examinations differ in frequency and time course across acute brain-injury phenotypes, and whether the handling convention changes illness-severity estimates and mortality-model behavior, has not been characterized at scale, even though every GCS-based ICU metric and risk-adjusted benchmark inherits this convention. We used a large critical care database to quantify the burden and temporal dynamics of non-assessable verbal examinations across six acute brain-injury phenotypes, to audit how commonly used handling strategies affect computed severity and an in-hospital-mortality model, and to test whether a sedation-aware machine-learning model can recover the non-assessable verbal component more accurately than the standard heuristic. To test whether the distortion is a general property of ICU scoring rather than a single-database artifact, we then replicated the two transportable claims in the multi-hospital eICU Collaborative Research Database and its APACHE physiology records.

## Methods

### Study Design and Setting

This was a retrospective cohort study of MIMIC-IV version 3.1, a single-center critical care database of adults admitted to the intensive care units (ICUs) of Beth Israel Deaconess Medical Center between 2008 and 2019.^9^ Reporting followed STROBE and RECORD, and the prediction components followed TRIPOD where applicable.^8,10,11^ The four prespecified aims were the burden of non-assessable verbal examinations (Aim 1), their temporal dynamics and proximate determinants (Aim 2), an audit of how the handling of non-assessable examinations affects severity scoring and mortality-model behavior (Aim 3), and a secondary, exploratory test of whether a sedation-aware machine-learning model can recover the verbal component (Aim 4). No causal or treatment-effect claims were made; all analyses were descriptive or methodological.

### Cohort Identification

Adults 18 years or older with at least one ICU stay were eligible. Acute brain injury was defined from ICD-9 and ICD-10 diagnosis codes in any position (eTable 1), grouped into six phenotypes: traumatic brain injury, intracerebral hemorrhage, acute ischemic stroke, subarachnoid hemorrhage, subdural hemorrhage, and anoxic injury; when more than one was present, a single phenotype was assigned by a fixed clinical priority hierarchy (subarachnoid, then intracerebral, subdural, traumatic, ischemic, anoxic). A general-ICU comparator comprised stays without any brain-injury diagnosis. The analytic cohort was the first ICU stay of each hospitalization with at least one charted Glasgow Coma Scale (GCS) verbal-component entry.

### Data Sources and Variables

The Glasgow Coma Scale components were extracted from nursing flowsheets (eye 220739, verbal 223900, motor 223901). The verbal component was the index measurement, because in MIMIC-IV it carries an explicit token, “No Response-ETT,” recording that the patient could not be assessed verbally owing to an endotracheal or tracheostomy tube. A verbal entry was non-assessable when its value was “No Response-ETT”; this is a documented inability to assess, not a verbal score of 1 (scoring it 1 is one handling strategy evaluated, not the study definition; value set, eTable 2). A sensitivity definition broadening non-assessability to documented intubation with a null or “No Response” entry was prespecified, and periods of active neuromuscular blockade were derived from infusion records.

Sedation depth (Richmond Agitation-Sedation Scale), continuous sedative, analgesic, neuromuscular-blocking, and vasopressor infusions, and invasive mechanical ventilation episodes were extracted as timestamped events; demographic characteristics, hospital discharge disposition, and the in-hospital mortality flag were taken from the admissions and patients tables (item identifiers in eTable 5).

### Outcomes and Measures

For Aim 1, burden was the proportion with any non-assessable verbal examination and the per-stay non-assessable fraction, overall and within the first 24, 48, and 72 hours, for the whole phenotype and the ventilated subgroup. For Aim 2, temporal dynamics were the time to the first non-assessable examination and a population assessability curve (fraction assessable per hour, by phenotype); determinants were the proportion of non-assessable examinations coinciding within 2 hours with a sedative, neuromuscular-blocking, or ventilation episode.

For Aim 3, the outcome was in-hospital mortality. Four strategies that are used in the published critical care modeling literature to handle a non-assessable verbal component were compared: (1) complete-case analysis, in which patients whose worst first-day examination was non-assessable were excluded; (2) fixed imputation of the verbal component to 1; (3) reporting the eye-plus-motor sum with a “T” designation; and (4) the eye-and-motor-based verbal imputation of Brennan et al (the exact eye-and-motor-to-verbal mapping is in eTable 8). Strategies were applied to the worst (lowest eye-plus-motor) GCS observation in the first 24 hours of each stay. We also reconstructed the GCS as produced by the official MIMIC-code derivation (gcs.sql), which sets the total GCS to 15 when the verbal entry is “No Response-ETT”^17^, and compared it with a component-aware GCS (eye and motor plus the Brennan-imputed verbal) at the first-day minimum. Severity was categorized by the Sequential Organ Failure Assessment (SOFA) central nervous system score and the APACHE II GCS contribution; the derived GCS was also entered as a strategy in the mortality model.

For Aim 4, a secondary exploratory analysis, a sedation-aware gradient-boosted classifier was trained to recover the verbal component from variables available at the examination (eye and motor scores, nearest Richmond Agitation-Sedation Scale value, indicators for active sedative, neuromuscular-blocking, and vasopressor infusions, and age), using only assessable examinations in the first 72 hours, with recovery estimated by patient-level grouped 5-fold cross-validation. The mechanical-ventilation indicator was excluded because it is near-deterministic for non-assessability (probability 0.93 given active ventilation) yet nearly absent from the assessable training examinations (1.4% ventilated), so retaining it would risk circularity; a prespecified ablation confirmed it changed neither recovery nor discrimination (eTable 13). Recovery was benchmarked against the eye-and-motor heuristic and fixed imputation, and the learned model was added as a fifth handling strategy in the Aim 3 mortality model. A motor-component-only severity descriptor, which never requires the verbal component, was also compared with the eye-and-motor (Brennan) total (eTable 14).

### Statistical Analysis

Aims 1 and 2 were descriptive. Phenotype differences in the presence of any non-assessable verbal examination were tested with the χ² test (Cramér V effect size), and differences in the per-stay non-assessable fraction (ventilated patients) and the time to the first non-assessable verbal examination with the Kruskal-Wallis test (epsilon-squared).

The independent association of phenotype with ever being non-assessable was estimated with logistic regression adjusted for age and sex (acute ischemic stroke reference; mechanical ventilation, a mediator, excluded), reported as odds ratios with 95% confidence intervals. Predictors of a non-assessable worst examination (phenotype, age, mechanical ventilation, sedative and vasopressor infusions, and in-hospital mortality) were estimated with logistic regression, and standardized mean differences compared the complete-case excluded and retained patients.

For Aim 3, a deliberately parsimonious logistic model, intended as an audit instrument rather than a clinical prediction model, related in-hospital mortality to the total GCS (per strategy), age, vasopressor use, and mechanical ventilation; discrimination was estimated by 5-fold cross-validation as the area under the receiver operating characteristic curve (AUROC; bootstrap 95% confidence interval), Brier score, and calibration (eTable 9), overall and by phenotype, with paired-bootstrap comparison of strategies. The selection effect of complete-case handling was summarized by the relative risk and risk difference of mortality between excluded and retained patients, and reclassification by the proportion changing predicted-risk tertile. For Aim 4, verbal-score recovery was estimated by patient-level grouped 5-fold cross-validation as accuracy and the quadratic weighted kappa. The four primary tests were corrected with the Benjamini-Hochberg false discovery rate; the threshold was P < .05, two-sided. Full detail is in the eMethods; analyses used Python 3.9, and code is at https://github.com/Alon-Gorenshtein/The_unexaminable_brain.

### External Validation (eICU-CRD)

To test whether the two transportable claims, the default-to-normal convention and the complete-case selection trap, generalize beyond a single center, we replicated them in the eICU Collaborative Research Database version 2.0, a multi-hospital critical care database drawn from intensive care units across the United States.^18^ The analytic set comprised 171,177 ICU stays with both an Acute Physiology and Chronic Health Evaluation (APACHE) physiology record and a patient record. GCS components (eyes, motor, verbal) and an intubated flag were taken from the apacheApsVar table, and in-hospital mortality from the patient table. eICU does not carry a distinct non-assessable verbal token analogous to the MIMIC-IV “No Response-ETT” entry; its nurseCharting verbal GCS is stored only as a number from 1 to 5. We therefore used the APACHE components and the intubated flag, defining a stay as non-assessable when an APACHE GCS component was coded -1. For the default-to-normal analysis we computed, among intubated stays, the proportion carrying a total GCS of 15 and the proportion carrying a verbal score of 5 (“oriented”), neither of which a truly intubated patient can produce; non-intubated stays served as a reference. For the selection-trap analysis we compared in-hospital mortality between non-assessable and assessable stays. Both analyses were run in the full ICU population and in a neurological or cardiac-arrest admission-diagnosis subgroup, with proportions reported as percentages and 95% confidence intervals (Wilson method).

### Ethics

MIMIC-IV contains de-identified data maintained under a data use agreement overseen by the institutional review boards of the Massachusetts Institute of Technology and Beth Israel Deaconess Medical Center, which granted a waiver of informed consent for the original collection; access was through credentialed PhysioNet authorization. eICU-CRD is similarly de-identified and accessed under credentialed PhysioNet authorization. This secondary analysis of publicly available de-identified databases required no additional institutional review board approval.

## Results

### Cohort

Of 94,458 ICU stays in MIMIC-IV, 14,230 first ICU stays of adults with an acute brain injury and at least one charted Glasgow Coma Scale verbal-component entry formed the analytic cohort; 70,705 stays without a brain-injury diagnosis served as the comparator (eFigure 1). Median age was 67 years (IQR 54–79); 44.3% were women and 37.9% mechanically ventilated. In-hospital mortality ranged from 12.3% (traumatic brain injury) to 41.3% (anoxic injury), versus 9.4% in the comparator (Table 1).

**Figure 1.**
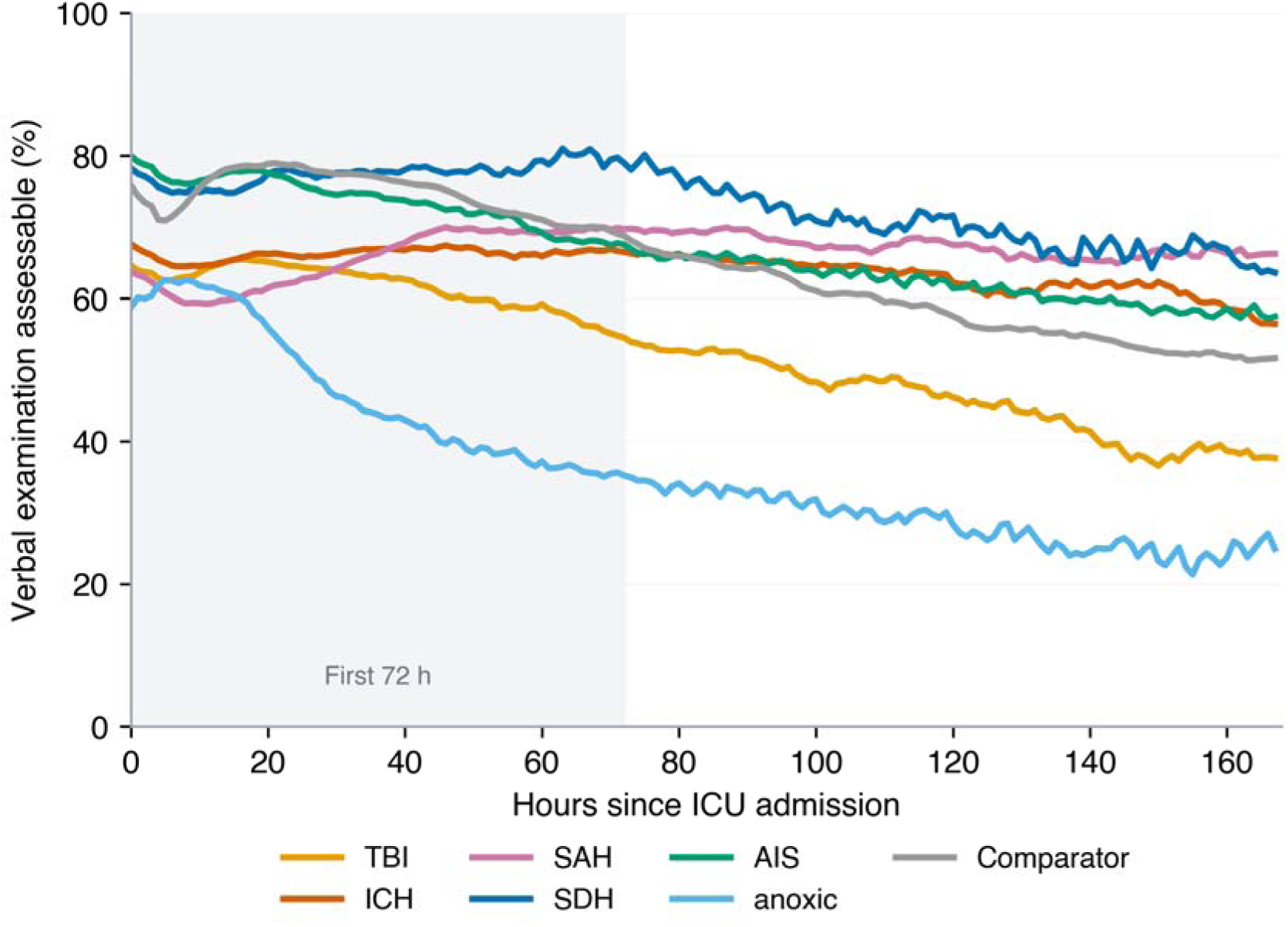
Population assessability of the verbal examination, by phenotype.

**Table 1.** Characteristics of the analytic cohort (first ICU stay with at least one charted GCS verbal-component entry), by acute brain-injury phenotype. Values are n (%) unless otherwise stated. TBI, traumatic brain injury; ICH, intracerebral hemorrhage; SAH, subarachnoid hemorrhage; SDH, subdural hemorrhage; AIS, acute ischemic stroke; LOS, length of stay.

| Characteristic | TBI | ICH | SAH | SDH | AIS | Anoxic | Comparator | Overall neuro |
| --- | --- | --- | --- | --- | --- | --- | --- | --- |
| N | 3874 | 2863 | 1314 | 804 | 4093 | 1282 | 70,705 | 14,230 |
| Age, median (IQR) | 65 (47-80) | 69 (57-80) | 61 (50-72) | 70 (59-79) | 70 (60-80) | 59 (47-69) | 64 (53-75) | 67 (54-79) |
| Female | 1446 (37.3) | 1299 (45.4) | 741 (56.4) | 295 (36.7) | 1967 (48.1) | 558 (43.5) | 31,273 (44.2) | 6306 (44.3) |
| White | 2474 (63.9) | 1753 (61.2) | 772 (58.8) | 532 (66.2) | 2635 (64.4) | 740 (57.7) | 47,448 (67.1) | 8906 (62.6) |
| Black | 202 (5.2) | 275 (9.6) | 99 (7.5) | 85 (10.6) | 413 (10.1) | 138 (10.8) | 8119 (11.5) | 1212 (8.5) |
| Medicare/Medic aid | 2417 (62.4) | 2028 (70.8) | 717 (54.6) | 590 (73.4) | 3057 (74.7) | 793 (61.9) | 49,613 (70.2) | 9602 (67.5) |
| Mechanically ventilated | 1445 (37.3) | 1023 (35.7) | 586 (44.6) | 242 (30.1) | 1399 (34.2) | 705 (55.0) | 23,447 (33.2) | 5400 (37.9) |
| Hospital mortality | 475 (12.3) | 648 (22.6) | 306 (23.3) | 138 (17.2) | 624 (15.2) | 529 (41.3) | 6619 (9.4) | 2720 (19.1) |
| ICU LOS, days, | 1.9 (1.0- | 3.0 (1.6- | 6.2 (2.2- | 2.5 (1.2- | 2.7 (1.4- | 2.9 (1.2- | 1.8 (1.1- | 2.7 (1.3- |
| median (IQR) | 4.3) | 6.8) | 12.8) | 4.9) | 5.5) | 6.0) | 3.3) | 6.0) |

### Burden of Non-Assessable Examinations (Aim 1)

Nearly half of brain-injured ICU patients had at least one verbal examination that could not be assessed. Across the neuro cohort, 45.2% had at least one non-assessable verbal examination versus 38.2% of comparators (Table 2), differing across phenotypes (χ = 332.4, P < .001; Cramér V, 0.15), highest in anoxic injury (66.1%) and lowest in subdural hemorrhage (35.3%).

**Table 2.**
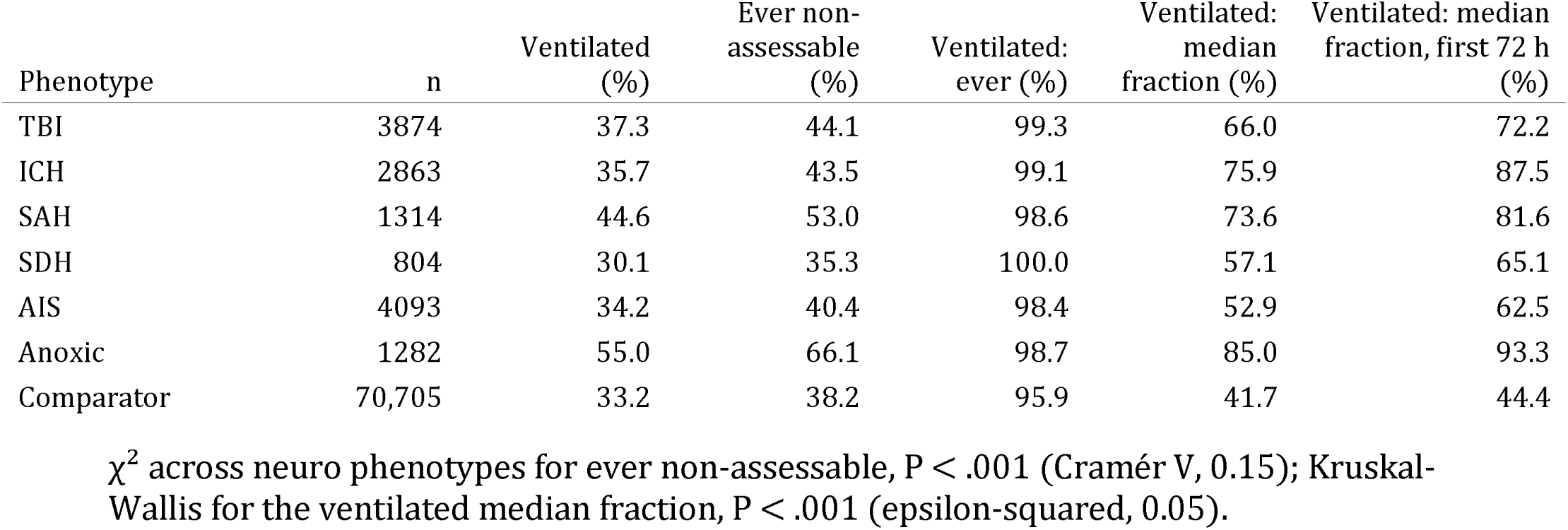
Burden of non-assessable neurological examinations, by phenotype (first ICU stay). “Ever non-assessable” is the percentage of patients with at least one non-assessable verbal examination. The ventilated columns are restricted to mechanically ventilated patients, in whom the phenomenon is concentrated. Median fraction is the per-patient percentage of verbal examinations recorded as non-assessable.

Among mechanically ventilated patients, non-assessable verbal examinations were near-universal and dominated the documented examination record. In the ventilated subgroup, 98.4% to 100% of patients had a non-assessable verbal examination, and the median patient had a majority of all verbal examinations recorded as non-assessable, ranging from 52.9% in acute ischemic stroke to 85.0% in anoxic brain injury (Kruskal-Wallis across phenotypes, H = 266.8, P < .001; epsilon-squared, 0.05; Table 2). Within the first 72 hours, the prognostically critical window, these fractions were higher still, reaching 93.3% in anoxic brain injury. Among ventilated patients, 47.5% had no assessable verbal examination at all during the first 24 hours, falling to 33.8% at 48 hours and 27.4% at 72 hours.

### Temporal Dynamics and Determinants (Aim 2)

The verbal examination went dark within the first hours of admission and stayed dark. Among patients who became non-assessable, the median time to the first non-assessable verbal examination was 0.9 to 1.3 hours in intracerebral hemorrhage, traumatic brain injury, subarachnoid hemorrhage, and anoxic injury (2.7 hours in ischemic stroke; 3.0 in the comparator; Kruskal-Wallis, H = 261.7, P < .001; epsilon-squared, 0.04). Population assessability curves remained below baseline throughout the first week and recovered slowly, with anoxic brain injury showing the lowest and most sustained loss of assessability (Figure 1).

Non-assessability tracked mechanical ventilation and sedation rather than neuromuscular blockade. Across phenotypes, 83% to 85% of non-assessable examinations coincided with mechanical ventilation and 59% to 70% with sedation, but under 9% with neuromuscular blockade (eFigure 2).

**Figure 2.**
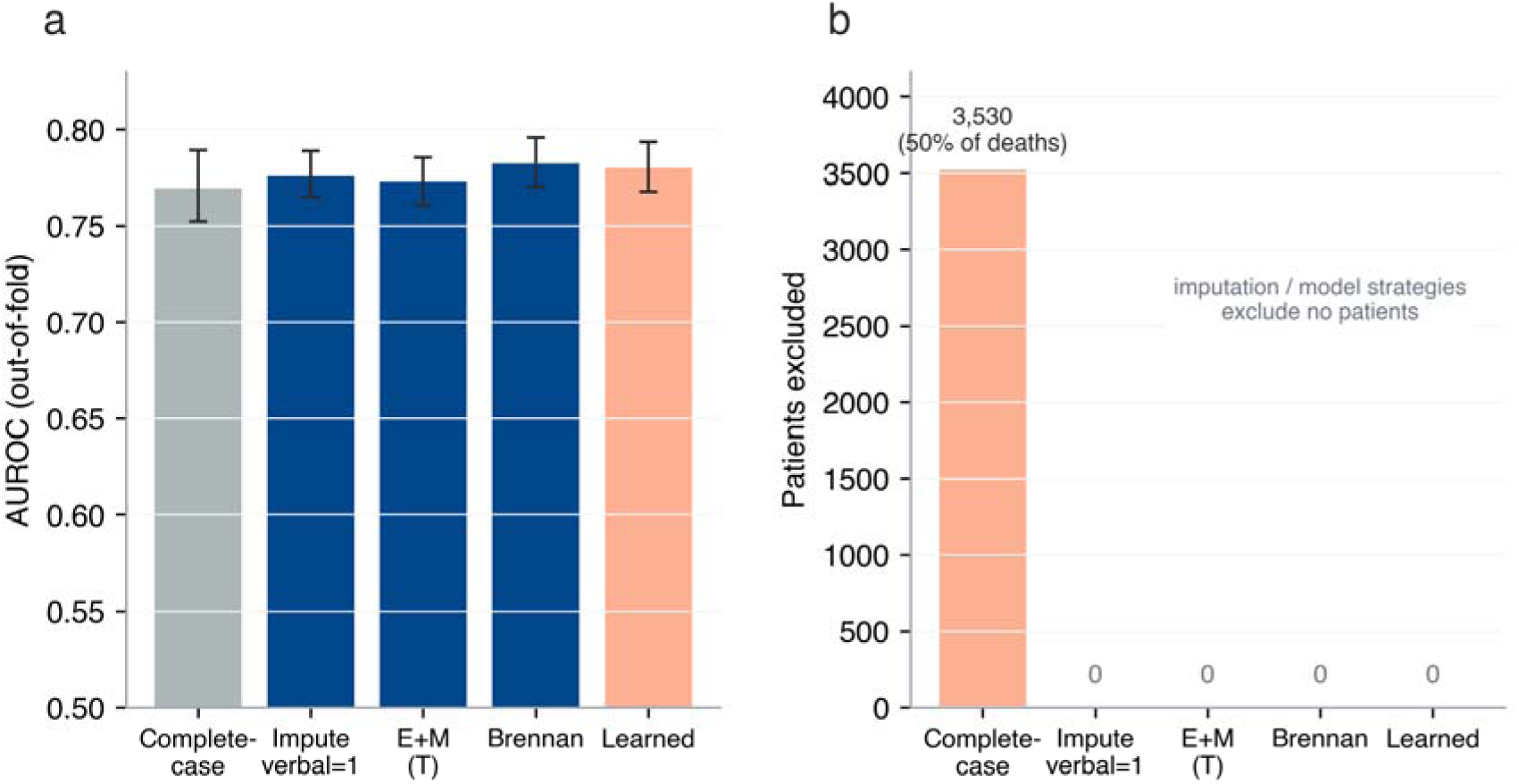
Computed total GCS by handling strategy.

Non-assessability was not missing at random. In a multivariable model, a non-assessable worst examination was driven by mechanical ventilation (odds ratio 27.4; 95% CI 22.8 to 33.0) and active sedation (odds ratio 2.54; 95% CI 2.11 to 3.06) and was independently associated with in-hospital mortality (odds ratio 1.56; 95% CI 1.32 to 1.85; eTable 11). Patients excluded by complete-case handling differed markedly from those retained, with large standardized mean differences for mechanical ventilation (2.32), sedation (1.83), and mortality (0.36; eTable 12).

### Analytic Consequences of Handling Choice (Aim 3)

The Aim 3 cohort comprised 12,404 patients with a complete worst first-day examination, of whom 3530 (28.5%) had a non-assessable verbal component.

A widely used derived-score convention normalized intubated patients to a perfect score. As a concrete example, the official MIMIC-code derivation assigned a first-day GCS of 15 to 5321 of 12,404 patients (42.9%); for 25.0% it overstated the component-aware GCS by a median of 5.5 points (IQR 2 to 8). It placed 11.6% in the lowest severity category (SOFA central nervous system score 0) although eye and motor components indicated GCS 9 or less, including 299 in deepest coma (Figure 3). The derived rule understated the APACHE II GCS contribution (1.58 vs 2.85 points) and discriminated mortality worse (AUROC 0.746; 95% CI 0.732 to 0.759 vs 0.783; Figure 3C).

**Figure 3.**
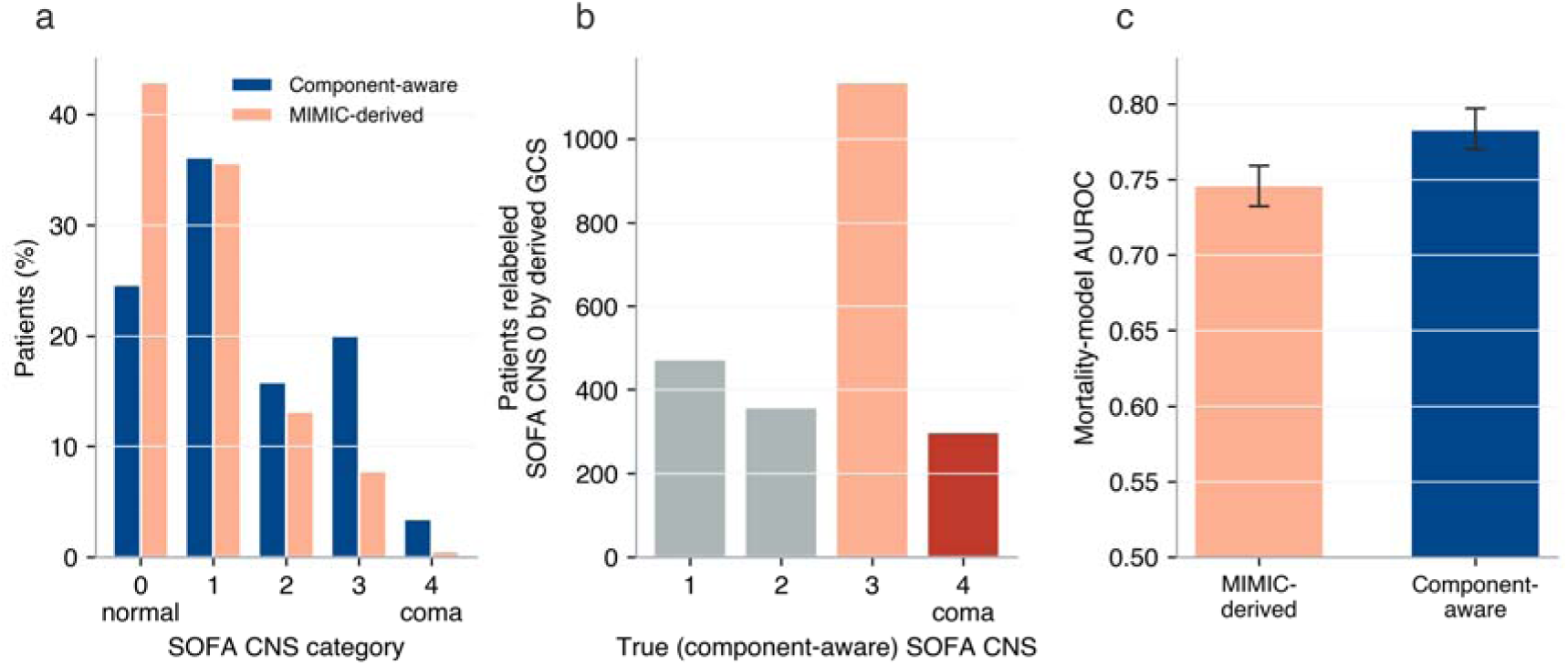
The MIMIC-derived GCS versus a component-aware GCS.

Complete-case handling silently discarded the highest-risk patients. Excluding patients whose worst verbal examination was non-assessable removed 3530 of 12,404 (28.5%). These excluded patients accounted for 720 of 1434 deaths (50.2%), with mortality 20.4% versus 8.0% among retained patients (relative risk, 2.54; 95% CI, 2.30 to 2.79; P < .001; Table 3), so the complete-case model was fit on a population with one-third the mortality of the full cohort.

**Table 3.**
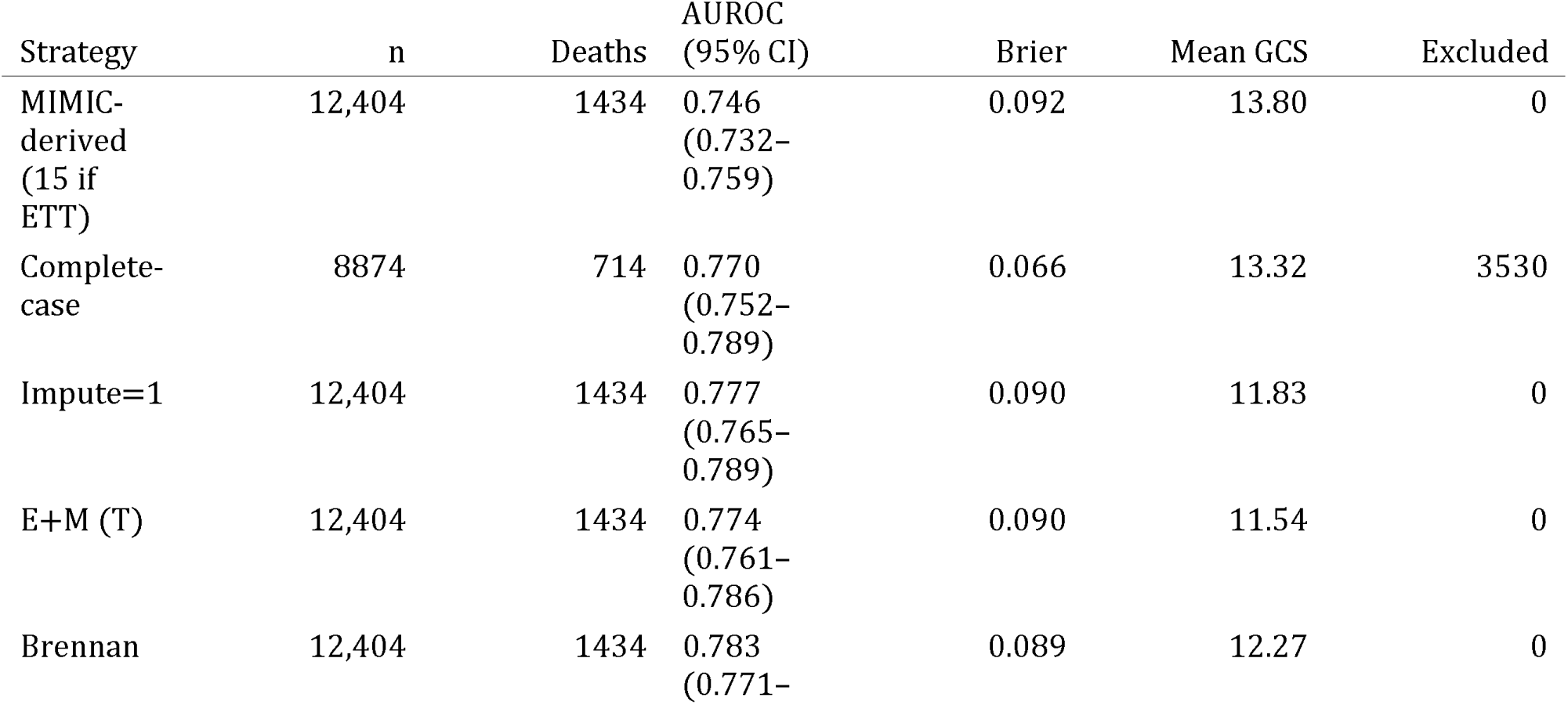
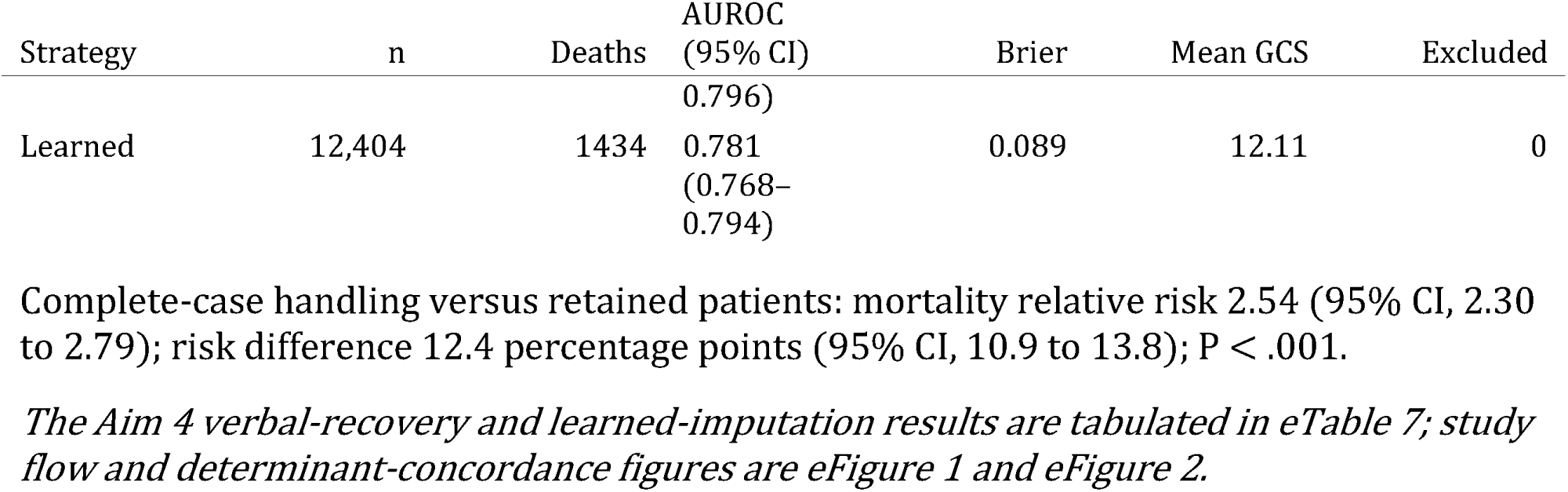
Effect of the GCS verbal-handling strategy on the in-hospital-mortality model (all neuro patients). MIMIC-derived applies the official gcs.sql rule (total GCS set to 15 when verbal is non-assessable). Complete-case excludes patients with a non-assessable verbal component; Impute=1 sets the verbal score to 1; E+M reports the eye-plus-motor sum; Brennan applies the eye-and-motor imputation; Learned applies the sedation-aware model. AUROC is out-of-fold with a bootstrap 95% CI. Per-phenotype results are in eTable 3.

The handling strategy changed the computed severity and the model’s risk estimates. Among the imputation strategies the mean total GCS ranged from 11.54 (eye-plus-motor convention) to 13.32 (complete-case), up to 1.8 points from the handling rule alone (Table 3, Figure 2), while discrimination was similar (AUROC 0.770 to 0.783). In the paired bootstrap the Brennan imputation discriminated marginally better (all P < .001) but by less than 0.02 AUROC, and 7.5% of patients changed predicted-risk tertile between fixed and Brennan imputation. Calibration, assessed on each strategy’s own cross-validated predictions, was near-ideal for every strategy (eTable 9) and did not distinguish them; the differences lay in the cohort and the individual risk estimates, not in a summary metric.

### Learned Recovery of the Verbal Examination (Aim 4)

In a secondary, exploratory analysis we tested whether a better imputation could recover the lost information rather than merely fill the gap. In patient-level cross-validation (240,159 examinations from 11,577 patients), a sedation-aware model recovered the verbal score more accurately than the eye-and-motor heuristic (quadratic weighted kappa 0.56; 95% CI 0.56 to 0.56 vs 0.41; accuracy 66.0% vs 61.0%) and far better than fixed imputation (kappa 0.00; eFigure 3, eTable 7). When used to impute the non-assessable component, however, it did not improve mortality discrimination (AUROC 0.781; 95% CI 0.768 to 0.794 vs 0.783 for the heuristic) and assigned a similar total score. A prespecified ablation restoring the mechanical-ventilation indicator left both recovery and discrimination unchanged, confirming ventilation was not driving the result (eTable 13). A motor-component-only descriptor, which never requires the verbal component, discriminated mortality almost as well as the eye-and-motor (Brennan) total (AUROC 0.778; 95% CI 0.765 to 0.790 vs 0.783; eTable 14), consistent with most prognostic information residing in the motor response. Better reconstruction of the verbal score therefore did not recover prognostic information.

### Sensitivity Analyses

The same ordering of mean total GCS and the same complete-case selection effect held in each phenotype-stratified model (eTable 3), and the phenotype gradient in non-assessability burden was preserved under the broadened non-assessability definition, which changed the burden estimates by less than one percentage point because the added category is rare (eTable 4).

### External Validation in a Multi-Hospital Cohort

Both transportable findings replicated in eICU-CRD, a multi-hospital database. Among 171,177 ICU stays with paired APACHE and patient records, the default-to-normal convention recurred: of 25,398 intubated stays, 25.2% (95% CI, 24.7 to 25.7) carried a total GCS of 15 and 26.8% (95% CI, 26.3 to 27.4) carried a verbal score of 5 (“oriented”), values a truly intubated patient cannot produce, whereas 67.7% of non-intubated stays carried a verbal score of 5. In the neurological or cardiac-arrest admission subgroup, 8.1% of intubated stays still carried a total GCS of 15. The selection trap also replicated: stays with a non-assessable GCS had higher in-hospital mortality than assessable stays in the full ICU population (16.6% [95% CI, 15.7 to 17.6] vs 9.2% [95% CI, 9.1 to 9.4]) and in the neurological or cardiac-arrest subgroup (38.0% [95% CI, 34.4 to 41.8] vs 16.0% [95% CI, 15.6 to 16.4]), so restricting to assessable stays, the implicit complete-case choice, again dropped the sickest patients. These contrasts parallel the single-center MIMIC findings, in which 45.2% of neuro-ICU stays were ever non-assessable on the verbal GCS and the complete-case strategy selected lower-mortality patients, indicating that the convention and the selection trap are database-general rather than a single-center artifact.

## Discussion

In a cohort of more than 14,000 brain-injured ICU patients, the verbal examination was non-assessable in nearly half, near-universally so under mechanical ventilation, and the convention used to handle it changed both the computed illness severity and the study population. The default-to-15 convention is a concrete example: it normalizes intubated patients to a perfect neurologic score, and in this cohort it relabeled one in nine patients from a severe or comatose component-aware examination to the lowest severity category, understated the APACHE II GCS contribution by almost half, and degraded a mortality model, so the recorded severity tracks a documentation rule rather than the patient’s neurology.

Non-assessability is not random: it is driven by intubation and sedation, which accompany the most severe injuries, so its absence is informative. This explains the selection effect of complete-case handling, which leaves a model fit on a population with one-third the mortality of the cohort it represents. The near-identical discrimination across strategies is the point, not a reassurance: a summary metric can stay stable while the patients, the computed severity, and the individual risk estimates all shift.

Prior work on the missing verbal examination concentrated on imputation within one population: an eye-and-motor rule was derived and validated, and an estimated total GCS retained prognostic value in intubated traumatic brain injury in this database^5,6^, while simulations show imputation choice can alter model performance^7,16^. We instead quantify how wide the gap is, when it opens, and how the conventions used in practice, including the derived default-to-15, reshape the population and the severity estimate. The Full Outline of UnResponsiveness score addresses the unscorable verbal component at the bedside but not how retrospective studies should treat examinations already recorded as non-assessable^4^.

In a secondary, exploratory analysis, a sedation-aware machine-learning model recovered the verbal component more accurately than the heuristic but did not improve mortality discrimination; better reconstruction of the verbal score did not recover prognostic information, and a motor-component-only descriptor that never needs the verbal score discriminated nearly as well. Because this model is necessarily trained on assessable examinations and applied to the intubated examinations it can never observe, it is a probe rather than a deployable tool; the result was robust to excluding or including the near-deterministic ventilation indicator. The missingness is informative and convention-dependent, and no imputation, however accurate, substitutes for an examinable patient.

These findings carry three implications for critical care. First, the illness-severity scores and risk-adjusted mortality models that embed the GCS, including APACHE and SOFA^12,13^, are sensitive to this documentation convention, not to physiology alone: complete-case handling silently removed a quarter of patients and half of the deaths, and the default-to-15 convention understated the APACHE II GCS contribution by almost half and placed one in nine patients in the lowest severity category despite a severe or comatose eye-and-motor examination. Second, ICU benchmarking and quality metrics that use the GCS should flag and report how the non-assessable verbal examination was handled, because that choice, not case mix alone, can move a unit’s computed severity and risk-adjusted mortality. Third, the quantified window during which the bedside examination is unavailable in ventilated brain-injured patients (non-assessable a median of about 1 hour after admission and persisting through the first 72 hours in most ventilated patients) is a concrete denominator for exam-independent monitoring, such as continuous electroencephalography^14^ and automated pupillometry^15^, in exactly the patients in whom the examination is most needed. That the default-to-normal convention and the complete-case selection trap both replicated in the multi-hospital eICU-CRD/APACHE cohort indicates this is a general property of ICU scoring, not a single-database artifact.

Box 1. Recommended reporting of the non-assessable verbal GCS in ICU severity-scoring, benchmarking, and research. (1) The GCS component values used and their source. (2) How a non-assessable verbal examination is identified in the ventilated patient (for example the “No Response-ETT” token), and that it is not a verbal score of 1. (3) The handling rule applied (exclusion, fixed imputation, eye-plus-motor sum, eye-and-motor imputation, or the default-to-15), stated explicitly. (4) Whether a derived or automated GCS pipeline was used and how it treats intubation and sedation. (5) The proportion of the cohort or cohort-of-comparison affected. (6) A sensitivity analysis under at least one alternative rule.

### Study Limitations

This study has limitations. It addresses the verbal component; the eye and motor components remain documentable, so the burden is verbal-specific rather than absence of the whole examination. First, the primary cohort is single-center, and documentation and case mix may differ elsewhere; phenotypes were assigned from diagnosis codes in any position, so the phenotype contrasts may partly reflect coding depth. We mitigated the single-center concern by externally replicating the two transportable claims in the multi-hospital eICU-CRD database, where both the default-to-normal convention and the complete-case selection trap recurred, indicating they are database-general rather than a single institution’s quirk; the burden, temporal, and modeling analyses that depend on the granular MIMIC-IV chart, however, remain single-center. The replication carries a caveat: eICU’s nurseCharting stores the verbal GCS only as a number from 1 to 5 with no distinct non-assessable token, so the exact MIMIC-IV “No Response-ETT” entry cannot be reproduced, and we relied instead on the APACHE components and the intubated flag. That the two most widely used public ICU databases encode non-assessability in different yet equally lossy ways is itself consistent with the central thesis that derived-score conventions, not the underlying clinical reality, drive the recorded value. Second, non-assessability was identified from a charted token (“No Response-ETT”) rather than a record of an attempted examination, so it may misclassify in either direction, though a broadened definition gave the same ordering. Third, MIMIC-IV has no structured functional outcome, so the mortality model is a measurement vehicle and the learned model a secondary, exploratory probe, neither externally validated; the model’s recovery accuracy was necessarily estimated on assessable examinations, which differ systematically from the intubated examinations to which it is applied and in which no ground-truth verbal score exists, so it indexes performance on assessable examinations rather than verified recovery. The mechanical-ventilation indicator was excluded to avoid encoding the target, and a prespecified ablation confirmed its inclusion changed neither recovery nor discrimination; this guards against circularity but does not overcome the absence of a ground truth in the intubated patient, the reason Aim 4 is exploratory. Fourth, determinants were limited to ventilation, sedation, and neuromuscular blockade, as imaging, notes, and quantitative neuromonitoring are absent. Fifth, the analysis was descriptive and not designed to estimate treatment effects.

### Future Directions

Our external replication in eICU-CRD confirmed the convention and the selection trap across many hospitals, but the phenotype gradient, the temporal dynamics, and the full handling-strategy comparison still rest on the granular MIMIC-IV chart; multicenter replication of those finer analyses would establish whether they too are stable across institutions, and a reporting standard for the non-assessable verbal examination would let readers and benchmarking bodies judge the comparability of GCS-based ICU severity and quality metrics. Linking the window of non-assessability to the yield of monitoring that does not require patient participation, such as continuous electroencephalography^14^ or automated pupillometry^15^, would test whether the blind spot can be narrowed.

## Conclusion

In critically ill adults with acute brain injury, the verbal examination was non-assessable for a large, phenotype-dependent share of the early ICU course, near-universally so in mechanically ventilated patients, and the convention chosen to handle these examinations changed both the study population and the computed illness severity. Because this distortion held across two independent databases including 171 hospitals in eICU-CRD, it is a general property of GCS-based ICU scoring. ICU severity scores, risk-adjusted mortality models, and GCS-based benchmarking or quality metrics should report and justify how the non-assessable verbal examination was handled, because that choice, not only physiology, shapes the result.

## Supporting information

full appendix

## Data Availability

All data produced are available online at https://github.com/Alon-Gorenshtein/The_unexaminable_brain

## Competing interests

The authors declare that they have no competing interests.

## Funding

None.

## Notes

### Competing Interest Statement

The authors have declared no competing interest.

