## Supplementary material for "Default Handling of the Non-Assessable Verbal Glasgow Coma Scale Misclassifies Illness Severity in Mechanically Ventilated Patients: A Retrospective Analysis": full appendix

### SUPPLEMENTARY INFORMATION

#### Derived Clinical-Score Conventions Encode Informative Missingness in Public Critical Care Databases: The Non-Assessable Verbal Glasgow Coma Scale in MIMIC-IV

**Authors:** Alon Gorenshtein, MD[1,2]; Yosef Adiniaev[2]; Mahmud Omar, MD[2,3]; Yiftach Barash, MD[2,4]; Eyal Klang, MD[2,4]; Oved Daniel, MD[5]

**Affiliations:**

1. Department of Neurology, Beth Israel Deaconess Medical Center, Harvard Medical School, Boston, MA, USA
2. BRIDGE GenAI Lab, Beth Israel Deaconess Medical Center, Boston, MA, USA
3. The Windreich Department of Artificial Intelligence and Human Health, Mount Sinai Medical Center, NY, USA
4. Department of Radiology, Beth Israel Deaconess Medical Center, Harvard Medical School, Boston, MA, USA
5. Neurology Division, Tel Aviv Sourasky University Medical Center, Tel Aviv, Israel

This Supplementary Information accompanies the main manuscript. It contains the operational definitions used to identify acute brain-injury phenotypes and non-assessable verbal examinations, the source variable identifiers, the per-phenotype results of the handling-strategy audit, and a sensitivity analysis under a broadened definition of non-assessability. All analyses used MIMIC-IV version 3.1.

**Contents**


- S1 Supplementary Methods
- S2 Supplementary Tables (S1 to S14)
- S3 Supplementary Figures
- S4 Code and Data Availability


---

### S1 Supplementary Methods

#### S1.1 Phenotype definitions and priority hierarchy

Each hospitalization was assigned to one of six acute brain-injury phenotypes using International Classification of Diseases, Ninth and Tenth Revision (ICD-9 and ICD-10), diagnosis codes (eTable 1). When a hospitalization carried codes for more than one phenotype, a single phenotype was assigned by a fixed clinical priority hierarchy, applied so that the more acute and more procedurally specific diagnosis took precedence: subarachnoid hemorrhage, then intracerebral hemorrhage, subdural hemorrhage, traumatic brain injury, acute ischemic stroke, and anoxic injury. Diagnosis codes recorded in any position on the hospitalization were eligible. ICU stays without any brain-injury code formed the general-ICU comparator.

#### S1.2 Definition of a non-assessable verbal examination

The Glasgow Coma Scale verbal component (chartevents item 223900) was the index measurement. In MIMIC-IV the verbal component is recorded as one of six text values: Oriented, Confused, Inappropriate

Words, Incomprehensible sounds, No Response, and No Response-ETT. The first five are assessable responses; the sixth, No Response-ETT, denotes that the patient could not be assessed verbally because of an endotracheal or tracheostomy tube. A verbal entry was classified as non-assessable when its value was No Response-ETT (eTable 2). The complete value set and the count of each value across the database are given in eTable 2.

A broadened sensitivity definition also counted a verbal value of No Response as non-assessable when it was recorded during an active invasive mechanical ventilation episode (eTable 4). Periods of active neuromuscular blockade, during which the motor component is also uninterpretable, were identified from continuous infusion records (eTable 5).

#### S1.3 Source variables

All measurements were drawn from structured MIMIC-IV tables. The item identifiers used for the neurological examination, sedation, ventilation, and infusion variables are listed in eTable 5. Mechanical ventilation episodes were defined from invasive-ventilation procedure events (item 225792). Sedative, analgesic, neuromuscular-blocking, and vasopressor exposures were defined as the timestamped infusion intervals recorded in the inputevents table.

#### S1.4 Handling-strategy audit model

The Aim 3 audit used the single worst (lowest eye-plus-motor) Glasgow Coma Scale observation recorded in the first 24 hours of each ICU stay, with the eye and motor components mapped to their standard numeric values and the verbal value retained as recorded. Four handling strategies were applied to the verbal component when it was non-assessable: complete-case exclusion of the patient, fixed imputation of the verbal score to 1, reporting the eye-plus-motor sum (the "T" convention), and the eye-and-motor-based imputation of Brennan and colleagues (which adds 1 point to an eye-plus-motor sum of 2 to 6, 2 points to a sum of 7, 4 points to a sum of 8 or 9, and 5 points to a sum of 10).

For each strategy, a logistic regression model predicted in-hospital mortality from the total Glasgow Coma Scale (computed under that strategy), age, vasopressor use, and invasive mechanical ventilation. Features were standardized within each training fold. Discrimination was estimated by 5-fold stratified cross-validation using out-of-fold predicted probabilities, summarized by the area under the receiver operating characteristic curve (AUROC) with a 500-resample percentile bootstrap 95% confidence interval, and by the Brier score. The analysis was repeated within each phenotype (eTable 3). Risk reclassification between the fixed-imputation and Brennan strategies was summarized as the proportion of patients who changed predicted-risk tertile.

#### S1.5 Sedation-aware learned imputation (Aim 4)

Aim 4 was a secondary, exploratory analysis. A gradient-boosted classifier (scikit-learn HistGradientBoostingClassifier; 300 boosting iterations, learning rate 0.08, maximum tree depth 6) was trained to predict the verbal Glasgow Coma Scale category from seven inputs available at the time of the examination: the eye and motor scores, the nearest Richmond Agitation-Sedation Scale value within 2 hours, indicators for active sedative, neuromuscular-blocking, and vasopressor infusions, and age. The invasive mechanical ventilation indicator was deliberately excluded. Ventilation is near-deterministic for non-assessable status (the probability of a non-assessable verbal entry given an active ventilation episode was 0.93, versus 0.04 without) and is nearly absent from the assessable examinations on which the model is trained (1.4% ventilated), whereas the intubated examinations to which the model is applied are predominantly ventilated (81% at the worst first-day examination); retaining the indicator would therefore risk encoding the target itself when the model is extrapolated to the intubated population. A pre-specified ablation refitting the model with the ventilation indicator restored is reported in eTable 13 and confirmed that it changed neither recovery (quadratic weighted kappa 0.561 without versus 0.562 with) nor downstream discrimination (area under the curve 0.781 versus 0.777). Training and evaluation used only assessable examinations recorded in the first 72 hours, where the verbal component is observed (240,159 examinations from 11,577 patients). Recovery was estimated by patient-level 5-fold cross-validation (StratifiedGroupKFold grouped on patient identifier, so that no patient contributed examinations to both the training and the evaluation fold) and reported as accuracy and quadratic weighted kappa against the observed verbal score, each with a 500-resample bootstrap 95% confidence interval, and compared on the same held-out examinations with the eye-and-motor heuristic (whose implied verbal score is the heuristic addition) and with fixed imputation to 1. The model was then refit on all assessable examinations and used to impute the non-assessable verbal component, producing a fifth handling strategy for the Aim 3 mortality model. Agreement with the heuristic on non-assessable examinations was summarized as the difference in total Glasgow Coma Scale, overall and by sedation depth (eTable 6). As a further benchmark requested in review, a Glasgow Coma Scale motor-component-only severity descriptor, which never requires the verbal component, was entered into the Aim 3 mortality model and compared with the eye-and-motor (Brennan) total, overall and restricted to the non-assessable subset (eTable 14); the motor component was interpretable in essentially all non-assessable patients, with active neuromuscular blockade present in 0.1% of non-assessable worst examinations. The learned model is a measurement device used to probe the behavior of the conventions; it is not proposed as a deployable clinical tool, and because no ground-truth verbal score exists for the intubated examinations to which it is applied, its recovery accuracy is an upper bound estimated on assessable examinations rather than verified performance on the target.

##### S1.6 Software

Analyses were performed in Python 3.9 with pandas 2.3, NumPy 2.0, scikit-learn 1.6, statsmodels 0.14, SciPy 1.13, and matplotlib 3.9.

---

#### S2 Supplementary Tables

**eTable 1. Diagnosis codes used to define acute brain-injury phenotypes.** Codes are matched as prefixes on the recorded diagnosis code for the hospitalization.

| Phenotype | ICD-9 | ICD-10 |
|---|---|---|
| Subarachnoid hemorrhage | 430 | I60 |
| Intracerebral hemorrhage | 431 | I61 |
| Subdural hemorrhage | 432 | I62 |
| Acute ischemic stroke | 433, 434 | I63 |
| Traumatic brain injury | 800-804, 850-854 | S06 |
| Anoxic / hypoxic-ischemic injury | 348.1, 348.5 | G93.1, G93.82 |

**eTable 2. Recorded values of the Glasgow Coma Scale verbal component and their frequency across MIMIC-IV.** The value No Response-ETT denotes a non-assessable verbal examination owing to an endotracheal or tracheostomy tube.

| Verbal value | Assessable | Count |
|---|---|---|
| Oriented | Yes | 962,800 |
| No Response-ETT | No (non-assessable) | 784,911 |
| Confused | Yes | 273,419 |
| No Response | Yes | 103,056 |
| Incomprehensible sounds | Yes | 56,279 |
| Inappropriate Words | Yes | 24,656 |

**eTable 3. Handling-strategy audit, by phenotype.** For each phenotype and handling strategy, the analytic n, number of in-hospital deaths, out-of-fold AUROC with 95% CI, Brier score, mean total Glasgow Coma Scale, and number of patients excluded by the strategy. Complete-case is the only strategy that excludes patients.

| Phenotype | Strategy | n | Deaths | AUROC (95% CI) | Brier | Mean total GCS | Excluded |
|---|---|---|---|---|---|---|---|
| All neuro | Complete-case | 8,874 | 714 | 0.770 (0.752, 0.789) | 0.066 | 13.32 | 3,530 |
| All neuro | Impute verbal=1 | 12,404 | 1,434 | 0.777 (0.765, 0.789) | 0.090 | 11.83 | 0 |
| All neuro | E+M (T) | 12,404 | 1,434 | 0.774 (0.761, 0.786) | 0.090 | 11.54 | 0 |
| All neuro | Brennan | 12,404 | 1,434 | 0.783 (0.771, 0.796) | 0.089 | 12.27 | 0 |
| TBI | Complete-case | 2,401 | 113 | 0.842 (0.807, 0.881) | 0.040 | 13.54 | 1,084 |
| TBI | Brennan | 3,485 | 223 | 0.844 (0.824, 0.868) | 0.052 | 12.50 | 0 |
| ICH | Complete-case | 1,826 | 168 | 0.808 (0.771, 0.841) | 0.073 | 13.00 | 602 |
| ICH | Brennan | 2,428 | 324 | 0.800 (0.773, 0.824) | 0.098 | 12.12 | 0 |
| SAH | Complete-case | 742 | 54 | 0.761 (0.691, 0.827) | 0.061 | 13.31 | 340 |
| SAH | Brennan | 1,082 | 136 | 0.787 (0.742, 0.826) | 0.096 | 12.11 | 0 |

| Phenotype | Strategy | n | Deaths | AUROC (95% CI) | Brier | Mean total GCS | Excluded |
|---|---|---|---|---|---|---|---|
| SDH | Complete-case | 591 | 44 | 0.706 (0.604, 0.800) | 0.063 | 13.20 | 131 |
| SDH | Brennan | 722 | 73 | 0.754 (0.687, 0.825) | 0.079 | 12.49 | 0 |
| AIS | Complete-case | 2,784 | 266 | 0.720 (0.686, 0.753) | 0.080 | 13.41 | 1,024 |
| AIS | Brennan | 3,808 | 468 | 0.728 (0.702, 0.752) | 0.099 | 12.42 | 0 |
| Anoxic | Complete-case | 530 | 69 | 0.758 (0.684, 0.825) | 0.092 | 13.14 | 349 |
| Anoxic | Brennan | 879 | 210 | 0.797 (0.764, 0.830) | 0.142 | 11.11 | 0 |

In anoxic brain injury, complete-case handling excluded 349 of 879 patients (39.7%) and 141 of 210 deaths (67.1%). The impute-verbal=1 and E+M (T) rows for each phenotype share the same n and deaths as the Brennan row and are omitted here for brevity; full values are in the analysis output table.

**eTable 4. Sensitivity of the non-assessability burden to a broadened definition.** Percentage of patients ever non-assessable under the primary definition (No Response-ETT only) and under a broadened definition that also counts No Response recorded during active mechanical ventilation. The phenotype ordering is preserved.

| Phenotype | Ever non-assessable, primary (%) | Ever non-assessable, broadened (%) |
|---|---|---|
| TBI | 44.1 | 44.2 |
| ICH | 43.5 | 43.6 |
| SAH | 53.0 | 53.1 |
| SDH | 35.3 | 35.3 |
| AIS | 40.4 | 40.6 |
| Anoxic | 66.1 | 66.4 |
| Comparator | 38.2 | 38.7 |

**eTable 5. Source variable identifiers (MIMIC-IV).**

| Variable | Table | Item identifier(s) |
|---|---|---|
| GCS eye / verbal / motor | chartevents | 220739 / 223900 / 223901 |

| Variable | Table | Item identifier(s) |
|---|---|---|
| Richmond Agitation-Sedation Scale | chartevents | 228096 |
| Invasive mechanical ventilation | procedureevents | 225792 |
| Sedative and analgesic infusions | inputevents | 222168 (propofol), 221668 (midazolam), 225150 / 229420 (dexmedetomidine), 221744 / 225942 / 225972 (fentanyl), 221712 (ketamine), 221385 (lorazepam), 225156 (pentobarbital) |
| Neuromuscular blocking infusions | inputevents | 221555 (cisatracurium), 222062 (vecuronium), 229233 (rocuronium) |
| Vasopressor infusions | inputevents | 221906 (norepinephrine), 222315 (vasopressin), 221749 (phenylephrine), 221662 (dopamine), 221289 (epinephrine) |

**eTable 6. Divergence of the learned imputation from the eye-and-motor heuristic on non-assessable examinations, by sedation depth.** Difference is the learned total Glasgow Coma Scale minus the heuristic total. Negative values indicate the learned model assigned a lower score.

| Sedation depth | n | Mean difference | SD |
|---|---|---|---|
| Deep sedation (RASS <= -3) | 1418 | -0.66 | 1.05 |
| Light or none (RASS > -3) | 1602 | -0.39 | 1.23 |
| RASS missing | 510 | -0.63 | 1.31 |
| All non-assessable | 3530 | -0.54 | - |

Overall, the learned imputation differed from the heuristic in 46.9% of non-assessable examinations and was lower in 37.0%.

**eTable 7. Aim 4: recovery of the verbal component and its downstream effect.** Panel A reports recovery of the observed verbal score under patient-level (StratifiedGroupKFold) cross-validation, in which all of a patient's examinations are kept in the same fold. Panel B reports the in-hospital-mortality model when each method imputes the non-assessable component. QWK, quadratic weighted kappa.

*Panel A. Verbal-score recovery (n = 240,159 examinations from 11,577 patients)*

| Method | Accuracy (95% CI), % | QWK (95% CI) |
|---|---|---|
| Learned (sedation-aware) | 66.0 (65.8 to 66.2) | 0.56 (0.56 to 0.56) |
| Brennan (eye + motor) | 61.0 (60.8 to 61.2) | 0.41 (0.40 to 0.41) |
| Fixed verbal = 1 | 5.4 (5.3 to 5.5) | 0.00 |

The learned model used the seven-feature set without the mechanical-ventilation indicator (see S1.5 and eTable 13).

*Panel B. In-hospital-mortality model (n = 12,404)*

| Imputation | AUROC (95% CI) | Mean total GCS |
|---|---|---|
| Learned (sedation-aware) | 0.781 (0.768 to 0.794) | 12.11 |
| Brennan | 0.783 (0.773 to 0.796) | 12.27 |

**eTable 8. The Brennan eye-and-motor-to-verbal imputation.** When the verbal component is non-assessable, the eye-plus-motor sum determines the verbal points added to obtain the total GCS (Brennan et al, J Neurosurg 2020). The token "No Response-ETT" is treated as non-assessable, not as a verbal score of 1.

| Eye + motor sum | Imputed verbal points |
|---|---|
| 2 to 6 | +1 |
| 7 | +2 |
| 8 to 9 | +4 |
| 10 | +5 |

**eTable 9. Calibration of the in-hospital-mortality model by handling strategy (all neuro patients).** Calibration slope and calibration-in-the-large intercept from out-of-fold predictions. Because calibration was assessed on each model's own cross-validated predictions, all strategies were near-ideally calibrated by construction; the differences between strategies were in cohort composition and individual risk estimates.

| Strategy | n | Calibration slope | Calibration intercept |
|---|---|---|---|
| Complete-case | 8874 | 1.00 | 0.00 |
| Impute=1 | 12,404 | 1.00 | 0.00 |
| E+M (T) | 12,404 | 0.99 | 0.00 |
| Brennan | 12,404 | 1.00 | 0.00 |

**eTable 10. Raw component-aware versus official MIMIC-derived first-day GCS.** The MIMIC-code derivation (gcs.sql) sets the total GCS to 15 when the verbal entry is "No Response-ETT." Component-aware GCS uses the eye and motor components plus the Brennan-imputed verbal. SOFA, Sequential Organ Failure Assessment.

| Measure | Value |
|---|---|
| Patients with a derived first-day GCS of 15 | 5321 / 12,404 (42.9%) |
| Derived GCS overstated component-aware GCS | 3098 (25.0%); median +5.5 (IQR 2 to 8) points |
| Derived GCS 15 but component-aware GCS 8 or less | 845 (6.8%) |
| Derived SOFA CNS 0 but component-aware SOFA CNS 3 or worse | 1435 (11.6%) |

| Measure | Value |
|---|---|
| Mean APACHE II GCS contribution (derived vs component-aware) | 1.58 vs 2.85 |
| Mortality-model AUROC (derived vs component-aware) | 0.746 vs 0.783 |

SOFA central nervous system category cross-tabulation (rows, component-aware; columns, MIMIC-derived):

| Component-aware Derived | 0 | 1 | 2 | 3 | 4 |
|---|---|---|---|---|---|
| 0 | 3053 | 0 | 0 | 0 | 0 |
| 1 | 474 | 4009 | 0 | 0 | 0 |
| 2 | 359 | 143 | 1455 | 0 | 0 |
| 3 | 1136 | 239 | 166 | 941 | 0 |
| 4 (coma) | 299 | 25 | 12 | 25 | 68 |

**eTable 11. Predictors of a non-assessable worst first-day verbal examination (logistic regression).** Odds ratios with 95% confidence intervals; acute ischemic stroke is the phenotype reference.

| Predictor | OR (95% CI) | P |
|---|---|---|
| Mechanical ventilation | 27.41 (22.78 to 32.97) | <0.001 |
| Sedative infusion | 2.54 (2.11 to 3.06) | <0.001 |
| In-hospital mortality | 1.56 (1.32 to 1.85) | <0.001 |
| Vasopressor infusion | 0.76 (0.65 to 0.88) | <0.001 |
| Age (per year) | 0.99 (0.99 to 1.00) | <0.001 |
| Anoxic vs AIS | 1.37 (1.08 to 1.73) | 0.008 |
| TBI vs AIS | 1.24 (1.06 to 1.45) | 0.008 |
| SDH vs AIS | 0.56 (0.42 to 0.75) | <0.001 |

**eTable 12. Standardized mean differences between complete-case excluded and retained patients.** Excluded = non-assessable worst examination (n=3530); retained = assessable (n=8874). Standardized mean differences above 0.1 indicate imbalance.

| Variable | Excluded | Retained | SMD |
|---|---|---|---|
| Mechanically ventilated, % | 86.0 | 10.3 | 2.32 |
| Sedative infusion, % | 87.9 | 20.6 | 1.83 |
| Vasopressor infusion, % | 39.5 | 9.4 | 0.75 |
| Hospital mortality, % | 20.4 | 8.0 | 0.36 |
| Age, years | 61.3 | 66.6 | -0.30 |

**eTable 13. Aim 4 mechanical-ventilation ablation.** The reported sedation-aware verbal-recovery model excludes the mechanical-ventilation indicator because it is near-deterministic for non-assessable status and nearly absent from the assessable training examinations, so retaining it would risk circularity when the model

is applied to the predominantly ventilated intubated target. This ablation refits the model with the indicator restored. Recovery is the patient-level cross-validated quadratic weighted kappa and accuracy on assessable examinations (n = 240,159); downstream is the in-hospital-mortality model when the model imputes the non-assessable component (n = 12,404). Confidence intervals are 500-resample bootstrap percentiles. The two models are statistically indistinguishable, confirming the ventilation indicator carried no recovery signal.

| Model | Features | Recovery QWK (95% CI) | Recovery accuracy (95% CI), % | Downstream AUROC (95% CI) | Mean total GCS |
|---|---|---|---|---|---|
| Without ventilation flag (reported) | 7 | 0.561 (0.556 to 0.565) | 66.0 (65.8 to 66.2) | 0.781 (0.768 to 0.794) | 12.11 |
| With ventilation flag | 8 | 0.562 (0.558 to 0.566) | 66.2 (66.0 to 66.4) | 0.777 (0.764 to 0.790) | 11.97 |

In a permutation-importance analysis of the eight-feature model, the mechanical-ventilation indicator ranked sixth of eight (mean accuracy decrement 0.004), far below the motor score (0.069) and the Richmond Agitation-Sedation Scale value (0.046), consistent with its negligible contribution once eye, motor, and sedation depth are present.

**eTable 14. GCS-motor-only fallback benchmark (Aim 3 in-hospital-mortality model).** Some intubated patients retain an interpretable motor response. A motor-component-only severity descriptor (the motor score alone, which never requires the verbal component) was entered into the Aim 3 mortality model and compared with the eye-and-motor (Brennan) total. The motor component was interpretable in essentially all non-assessable patients (active neuromuscular blockade in 0.1% of non-assessable worst examinations). AUROC is out-of-fold with a 500-resample bootstrap 95% CI; the mean column reports the mean motor score for the motor-only rows and the mean total GCS for the Brennan reference.

| Scope | Descriptor | n | Deaths | AUROC (95% CI) | Brier | Mean |
|---|---|---|---|---|---|---|
| All neuro | GCS-motor-only | 12,404 | 1,434 | 0.778 (0.764 to 0.791) | 0.090 | 5.39 |
| All neuro | Brennan total (reference) | 12,404 | 1,434 | 0.783 (0.771 to 0.795) | 0.089 | 12.27 |
| Non-assessable only | GCS-motor-only | 3,530 | 720 | 0.746 (0.729 to 0.765) | 0.144 | 4.50 |

The motor-only descriptor discriminated in-hospital mortality almost as well as the full eye-and-motor total (difference in AUROC 0.005), indicating that most of the prognostic information attributed to the recovered verbal component in fact resides in the motor response. This supports the main finding that a more accurate verbal imputation does not recover prognostic information, and offers a transparent, imputation-free fallback for analyses of the intubated patient.

#### S3 Supplementary Figures

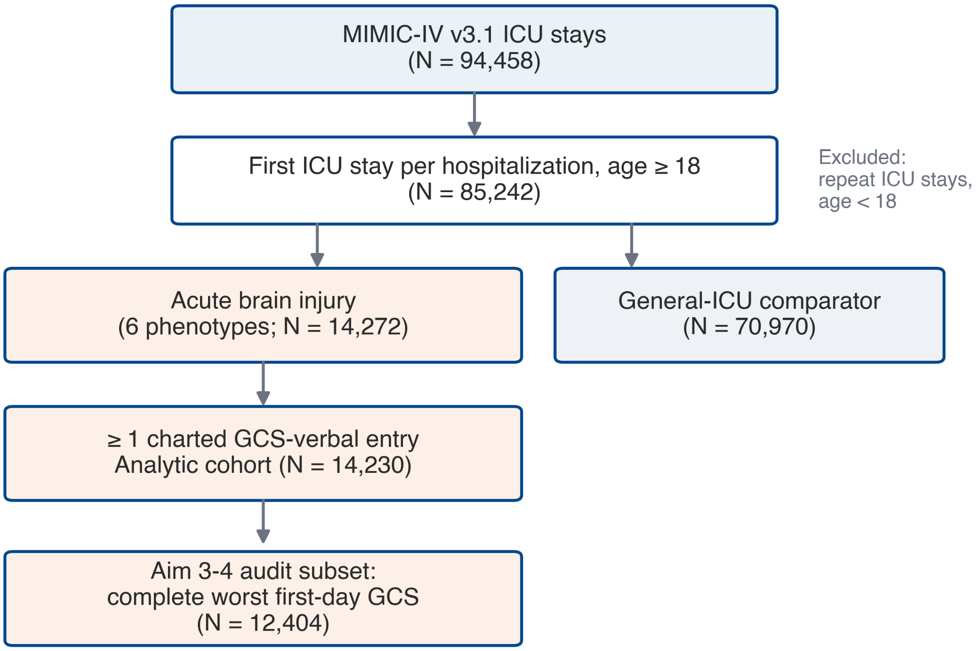


**eFigure 1. Study flow.** Derivation of the analytic cohort and the Aim 3-4 audit subset from MIMIC-IV.

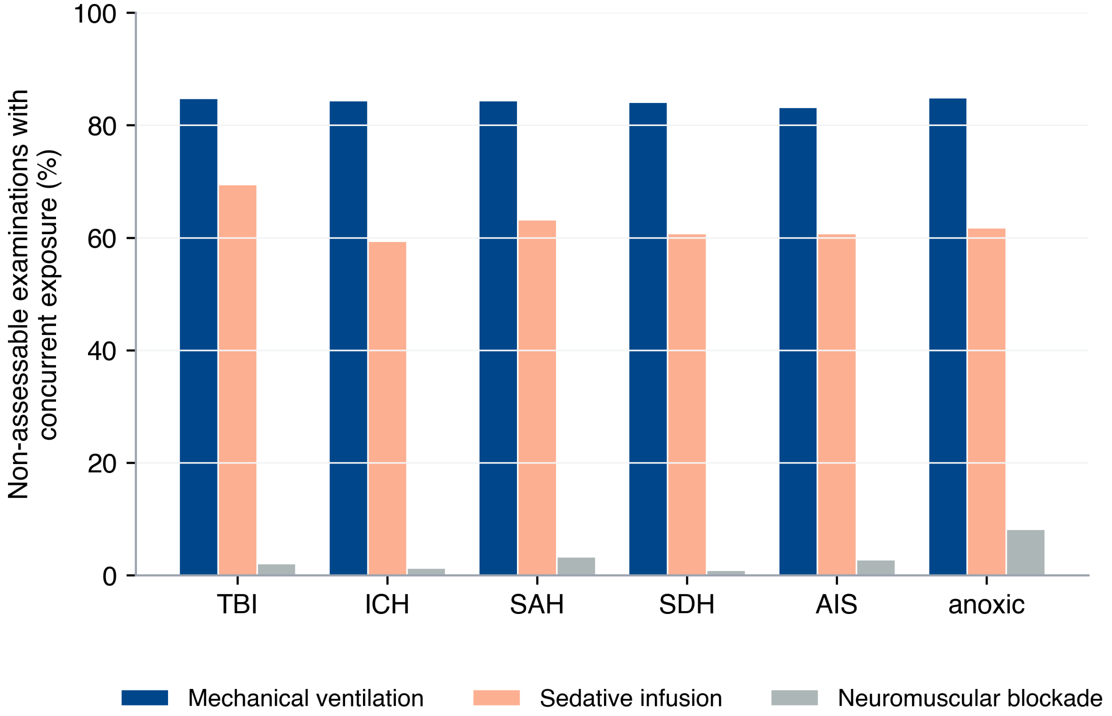


**eFigure 2. Determinants of non-assessability.** Proportion of non-assessable examinations coinciding with active mechanical ventilation, sedative infusion, or neuromuscular blockade, by phenotype.

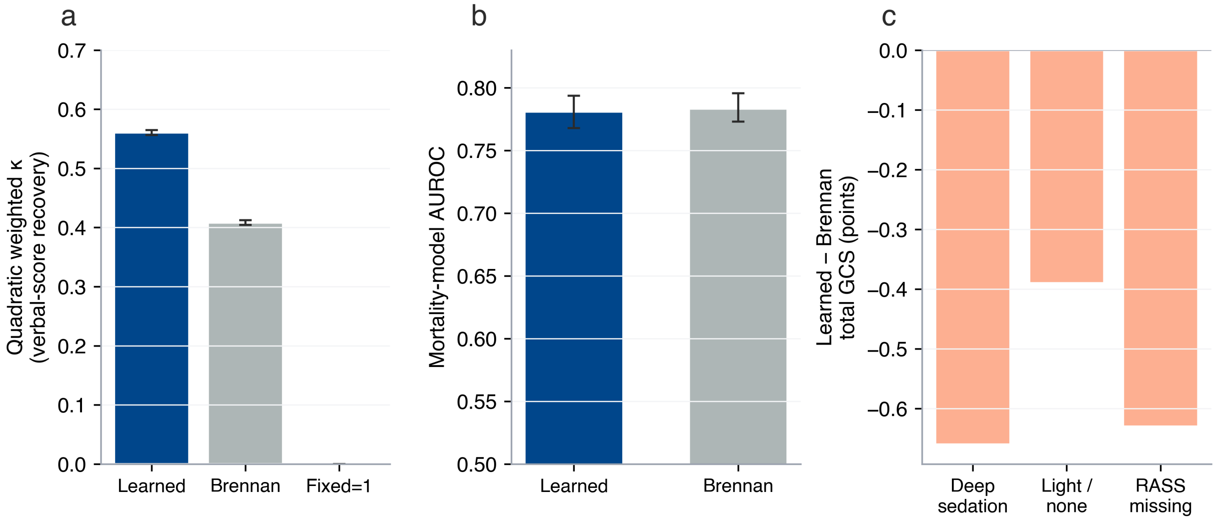


**eFigure 3. Sedation-aware learned imputation (Aim 4).** Verbal-score recovery under patient-level cross-validation (a), downstream mortality discrimination (b), and divergence of the learned imputation from the heuristic by sedation depth (c).

#### S4 Code and Data Availability

MIMIC-IV version 3.1 is available to credentialed users through PhysioNet under a data use agreement (https://physionet.org/content/mimiciv/3.1/). The analysis code that reproduces every table and figure from the source database is available at https://github.com/Alon-Gorenshtein/The_unexaminable_brain. No individual-level data are redistributed.
